# The spatial-temporal effect of air pollution on GP visits and hospital admissions by ethnicity in the United Kingdom: An individual-level analysis

**DOI:** 10.1101/2022.09.19.22280103

**Authors:** Mary Abed Al Ahad

## Abstract

**Background:** Air pollution has been associated with increased rates of hospital admissions and general-practitioner (GP) visits. Yet, more research is to be done to reveal the spatial-temporal dimension of this association and whether ethnic minorities experience greater effects from exposure to air pollution compared to the rest of population. This study investigates the spatial-temporal effect of air pollution on GP visits and hospital admissions by ethnicity in the United-Kingdom (UK).

**Methods:** We used individual-level longitudinal data from the *“UK Household Longitudinal Study”* including 46,442 adult individuals who provided 140,466 responses across five years (2015-2019). This data was linked to yearly concentrations of NO_2_, SO_2_, and particulate-matter (PM10, PM2.5) pollution using the Lower-Super-Output-Area (LSOA) of residence for each individual. We distinguished between spatial (*between* areas) and temporal (across time *within* each area) effects of air pollution on GP visits and hospital admissions and we used multilevel mixed-effects ordered logistic models for analysis.

**Results:** Results showed higher odds of outpatient hospital admissions with increasing concentrations of NO_2_ (OR=1.009; 95%CI=1.006-1.013), SO_2_ (OR=1.063; 95%CI=1.030-1.097), PM10 (OR=1.013; 95%CI=1.006-1.021), and PM2.5 (OR=1.022; 95%CI=1.012-1.032) pollutants. Higher odds of GP visits were also observed with increasing concentrations of NO_2_ (OR=1.011; 95%CI=1.007-1.015) and SO_2_ (OR=1.123; 95%CI=1.087-1.160) pollutants. Decomposing air pollution into *between* (spatial: across LSOAs) and *within* (temporal: across years within each LSOA) effects, showed significant *between* effects for air pollution on GP visits and hospital admissions, but not *within* effects. We observed no differences between ethnic minorities and British-white for the association between air pollution and hospital admissions and GP visits.

**Conclusion:** Using individual-level longitudinal data, our study supports the presence of a spatial-temporal association between air pollution and hospital admissions and GP visits. However, ethnic minorities do not seem to experience greater health-related effects from exposure to air pollution compared to the rest of population.

## 1. Introduction

Less than one year ago (31st October-12th November 2021), world leaders gathered at the COP26 in Glasgow-Scotland to tackle the issue of climate change with air pollution being one of its core elements and its consequences on humans’ health and planetary wellbeing. The association of air pollution with morbidity and mortality is well-documented in epidemiologic and environmental health studies which examine mild to severe outcomes including self-rated general health, doctor or general-practitioner (GP) visits, and hospital admissions, in particular those related to cancer, cardiovascular and respiratory health problems (1-4). Nevertheless, most of this research is concentrated on the health impacts of exposure to nitrogen dioxide (NO_2_), particulate matter with diameter of less than 10 μm (PM10) and particulate matter with diameter of less than 2.5 μm (PM2.5) pollution (2, 5). For example, in Lisbon-Portugal, higher rates of hospital admissions for circulatory and respiratory diseases were observed with higher concentrations of carbon monoxide, NO_2_, PM10 and PM2.5 pollutants (6). In New England, a 0.7% and a 4.2% increase in respiratory hospital admissions were noted for every 10 μg/m^3^ increase in short-term and long-term PM2.5 exposure, respectively (7). In Switzerland, doctor visits for chronic obstructive pulmonary diseases (COPD) and chronic bronchitis increased by 9.1% and 4.2%, respectively, with every 10 μg/m^3^ increase in NO_2_ pollutant (8). This highlights the need for studies that investigate the effect of other air pollutants (e.g. Sulphur dioxide [SO_2_]) on health-related outcomes (2).

Furthermore, the association between air pollution and health is not direct and several factors play a role in shaping this association. These factors include: (1) socioeconomic (e.g. age, gender, deprivation, income, socioeconomic classification, education, occupation, and economic activity) (2, 9-12); (2) individual and life-style (e.g. pre-existing diseases, cigarette smoking, alcohol drinking, physical exercise, and obesity) (13-16); (3) contextual (e.g. neighbourhood conditions, area-level multiple deprivation index, rural-urban classifications, and population density) (12, 17, 18); and (4) environmental (e.g. season, temperature, relative humidity, altitude, rainfall and wind) (19-21).

Despite the documented evidence about the factors affecting the association between air pollution and morbidity and mortality, published studies from Europe are mainly concentrated on assessing the differential effect of air pollution on health-related outcomes by gender, age, and socioeconomic status groupings, neglecting other key demographic sub-groups such as ethnicity (2). Ethnicity and country of birth are two important socio-demographic factors that affect individuals’ health (22, 23), and possibly lead to disparities in the effect of air pollution on hospital admissions and GP visits. Such ethnic disparities could be related to differences in the socioeconomic status (e.g. the relatively lower socioeconomic status for ethnic minorities), residential contexts (e.g. residing in more deprived neighbourhoods with inadequate housing conditions), exposure to ambient air pollution, or the differential effect of pollution by ethnicity (23-25). Thus, despite the “*Healthy migrant effect”*, which describes the advantageous characteristics (e.g. healthier, wealthier, and more educated) of immigrants (non-UK-born individuals) (26, 27), we expect ethnic minorities and non-UK-born individuals to show higher rates of hospital admissions and GP visits with increasing concentrations of air pollution. It should be noted, however, that literature on the effect of air pollution on health by ethnicity exist but is mainly focused around North America (28-30), which is characterised by a different ethnic composition to that in Europe.

In addition to the aforementioned literature gap, there is lack of research on the spatial-temporal effect of air pollution exposure on hospital admissions and GP visits using a *between-within* longitudinal design. The *between-within* design decomposes the overall effect of air pollution on hospital admissions and GP visits into *between* and *within* effects. The *Between* (spatial) effect assesses the effect of average air pollution concentrations across the follow-up time on hospital admissions and GP visits between geographical areas (e.g. census output areas) while the *within* effect (temporal) assesses the effect of the yearly deviation in air pollution concentrations from the average concentration on hospital admissions and GP visits across time within each geographical area (31). The benefits of the *between-within* approach is that it controls for data endogeneity and the effect of omitted confounders by allowing a time-varying geographical-fixed analysis (32); thus, providing detailed spatial-temporal evidence for policymaking decisions.

Considering the above-discussed literature gaps, this study utilises longitudinal individual-level data to investigate the spatial-temporal effect of air pollution on GP visits and hospital admissions by ethnicity in the UK. We aim specifically (1) to address the overall and the *between* (spatial-*between* geographical areas) and *within* (temporal-across time *within* each geographical area) effects of four air pollutants (NO_2_, SO_2_, PM10, and PM2.5) on hospital admissions and GP visits; and 2) to assess whether the association between air pollution and hospital admissions and GP visits varies between ethnic groups and by country of birth (non-UK-born versus UK-born).

## 2. Methods

### 2.1. Study design and population

We used longitudinal individual-level data from the “*Understanding Society: The UK Household Longitudinal Study (UKHLS)*” (33). This is a rich dataset, which consists of 10 data collection waves from 2009 to 2020 with around 40,000 households recruited at wave 1 from the four nations of the UK: England, Wales, Scotland, and Northern Ireland. It involves two main surveys: the youth survey which is filled by young people (aged 10 to 15) and the adult survey which is filled by individuals aged 16 and above (33).

The UKHLS dataset includes information on the socioeconomic characteristics of individuals including age, gender, marital status, education, occupation, socioeconomic classification, perceived financial situation, ethnicity, and country of birth and on the individuals’ self-rated health, wellbeing, hospital admissions, GP visits, smoking status, and the local authority and lower super output areas (LSOAs) of residence. Individuals recruited in the UKHLS are visited on a yearly basis to collect information on changes to their household and individual circumstances (33). Further information on the UKHLS dataset is described elsewhere (34-36).

For this study, we utilized individual-level data from the adult survey (age: 16+) of the UKHLS on 46,442 adult individuals who provided a total of 140,466 responses across four data collection waves (waves 7-10) over five years (2015-2019). We only used data from waves 7-10 of the UKHLS adult survey due to the unavailability of data on hospital admissions and GP visits in the earlier waves. We have to also note that the initial adult survey of the UKHLS for waves 7-10 involved a total of 49,028 individuals with 151,834 surveys and that 11,368 surveys were omitted due to the reasons described in Fig 1.

**Fig 1:**
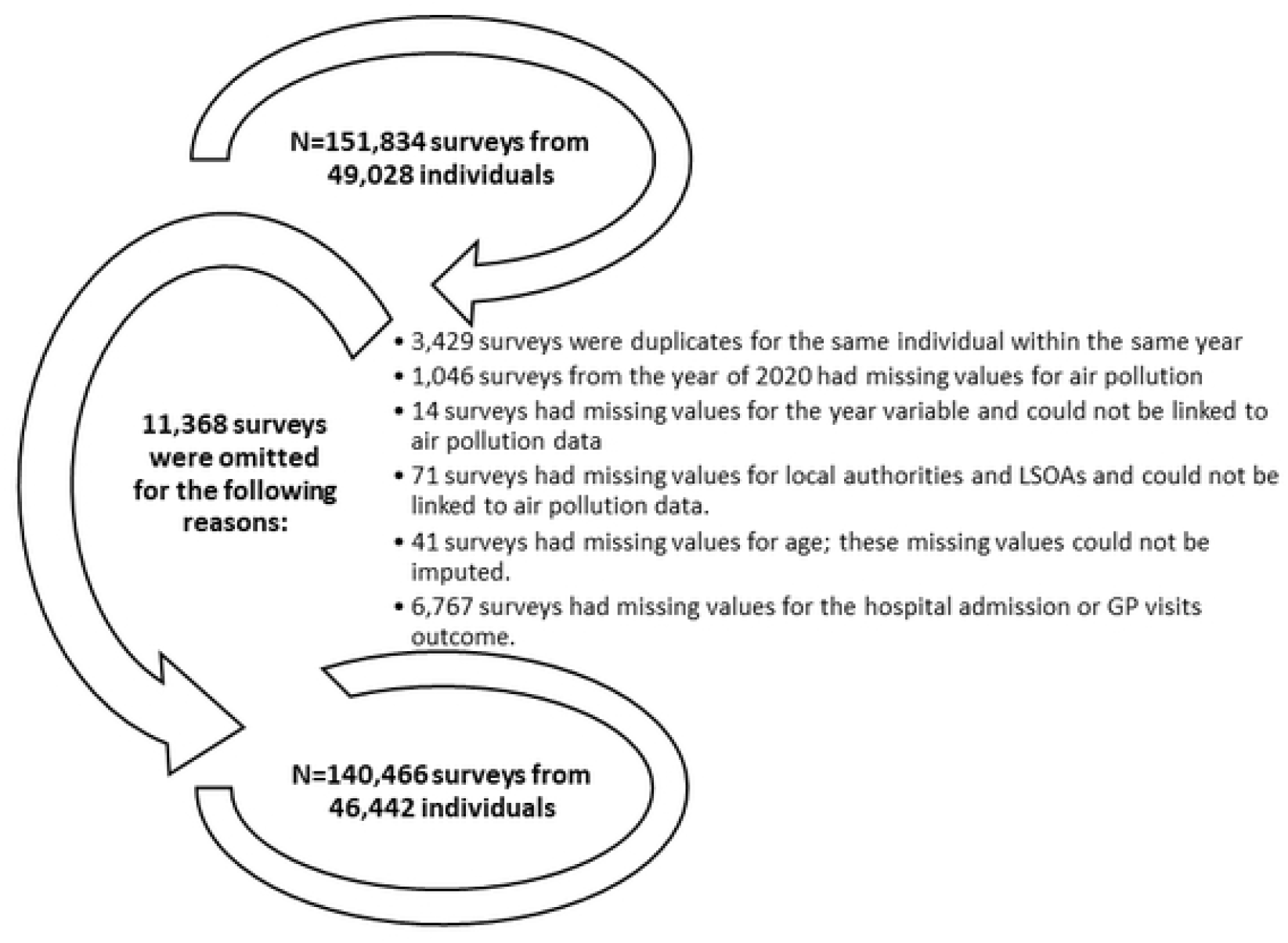
The reasons for omitting survey responses from the UKHLS (waves 7-10) data

### 2.2. Variables and measurements

#### 2.2.1. Outcome variables

*GP visits* and *outpatient hospital admissions* were measured as ordinal variables by asking individuals on the number of times they visited a GP or went to a hospital/clinic as outpatients in the past 12 months preceding the data collection date as follows: 0=none, 1=one to two times, 2=three to five times, 3=six to ten times, and 4=more than ten times.

#### 2.2.2. Air pollution

We downloaded yearly air pollution data that combine all sources of air pollution including road traffic and industrial/combustion processes for NO_2_, SO_2_, PM10, and PM2.5 pollutants from the “Department for Environment Food and Rural Affairs” online database (37). These are raster data of mean annual concentrations of pollutants measured in μg/m^3^ up to the year of 2019, estimated using air dispersion modelling (Pollution Climate Mapping) at a spatial resolution of 1×1 km and projected using the UK National Grid (37).

This 1×1 km raster air pollution data were linked to the UKHLS data using the Lower Super Output Areas (LSOAs; data zones for Scotland and Super Output Areas for Northern Ireland) of residence for each individual and each year (2015-2019). LSOAs are used to decompose England and Wales based on the population size into areas with a minimum population size of 1000 people and are the lowest level of geography offered by the UKHLS dataset. Some LSOAs, especially those located in urbanised areas, have an area of less than 1×1 km, which offers high spatial resolution to assess the impact of air pollution on hospital admissions and GP visits. The LSOAs in England and Wales are equivalent to data zones in Scotland and to Super Output Areas in Northern Ireland. For simplicity we refer to the joint LSOAs, data zones, and Super Output Areas as LSOAs. The linkage was done by computing the average air pollution concentration for each LSOA from all the 1×1 km raster squares that intersected with the respective LSOA. For example, if a certain LSOA intersected with only one raster square, it is given the value of air pollution of that square and if a certain LSOA intersected with two raster squares, it is given the average air pollution of those two squares, and so on so forth.

#### 2.2.3. Socioeconomic and lifestyle covariates

For this study, we selected a number of individual-level socioeconomic and lifestyle covariates based on what is available in the UKHLS dataset and based on the confounders and effect modifiers considered by relevant literature (1, 2). These included gender (1=male; 2=female); age (coded as 16-18 and then in 5 years increments as 19-23; 24-28; 29-33; 34-38; 39-43; 44-48; 49-53; 54-58; 59-63; 59-63; 64-68; 69-73; 74-78; >78); ethnicity (1=British-white; 2=Other-white; 3=Indian; 4=Pakistani/Bangladeshi; 5=Black/African/Caribbean; 6=mixed ethnicities; 7=Other ethnicities); country of birth (1=born in UK; 2=not born in UK; 3=No answer); marital status (1=married; 2=living as a couple; 3=widowed; 4=divorced/separated; 5=single never married; 6=no answer); education (1=university degree; 2=high school degree; 3=lower educational levels; 4=other qualifications; 5=still a student); perceived financial situation (1=living comfortably/doing alright; 2=living difficultly; 3=no answer); socioeconomic classification (1=management and professionals occupations; 2=intermediate occupations; 3=routine occupations; 4=not applicable: Student/retired/Not working; 5=no answer); and smoking status (0=non-smoker; 1=smoker; 2=no answer).

Finally, year dummies (calendar year: 2015-2019) were considered as a control for the time trend in our analysis following the approach of relevant studies (38, 39). Given that our study utilises yearly air pollution data, controlling for other temporal covariates considered by relevant literature such as seasonal trends (40, 41) was not possible.

### 2.3. Data analysis

Percentages were computed to describe the GP visits, hospital admissions, and individuals’ socioeconomic and lifestyle covariates for each wave (waves 7-10) of the UKHLS sample. We also examined the correlation between NO_2_, SO_2_, PM10, and PM2.5 pollutants at the LSOAs level using Pearson’s correlation coefficient. Given the high observed correlations between the pollutants (Pearson’s coefficient ≥ 0.7 (42); Table 2), the association of NO_2_, SO_2_, PM10, and PM2.5 pollutants with GP visits and hospital admissions was examined in separate regression models.

**Table 1:**
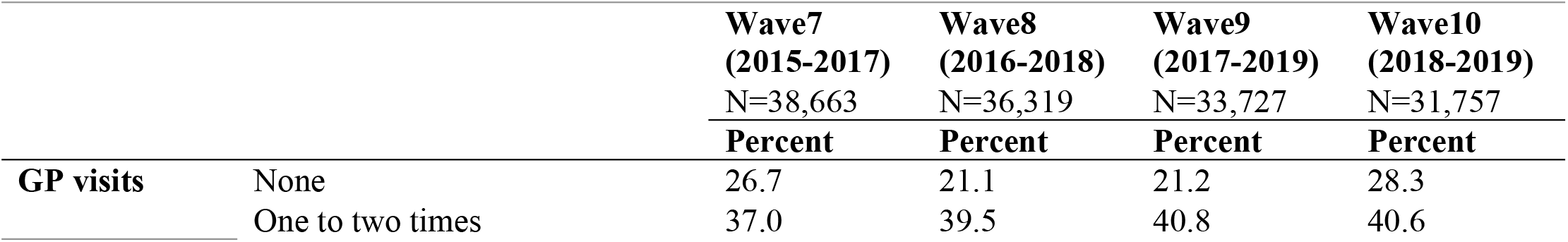
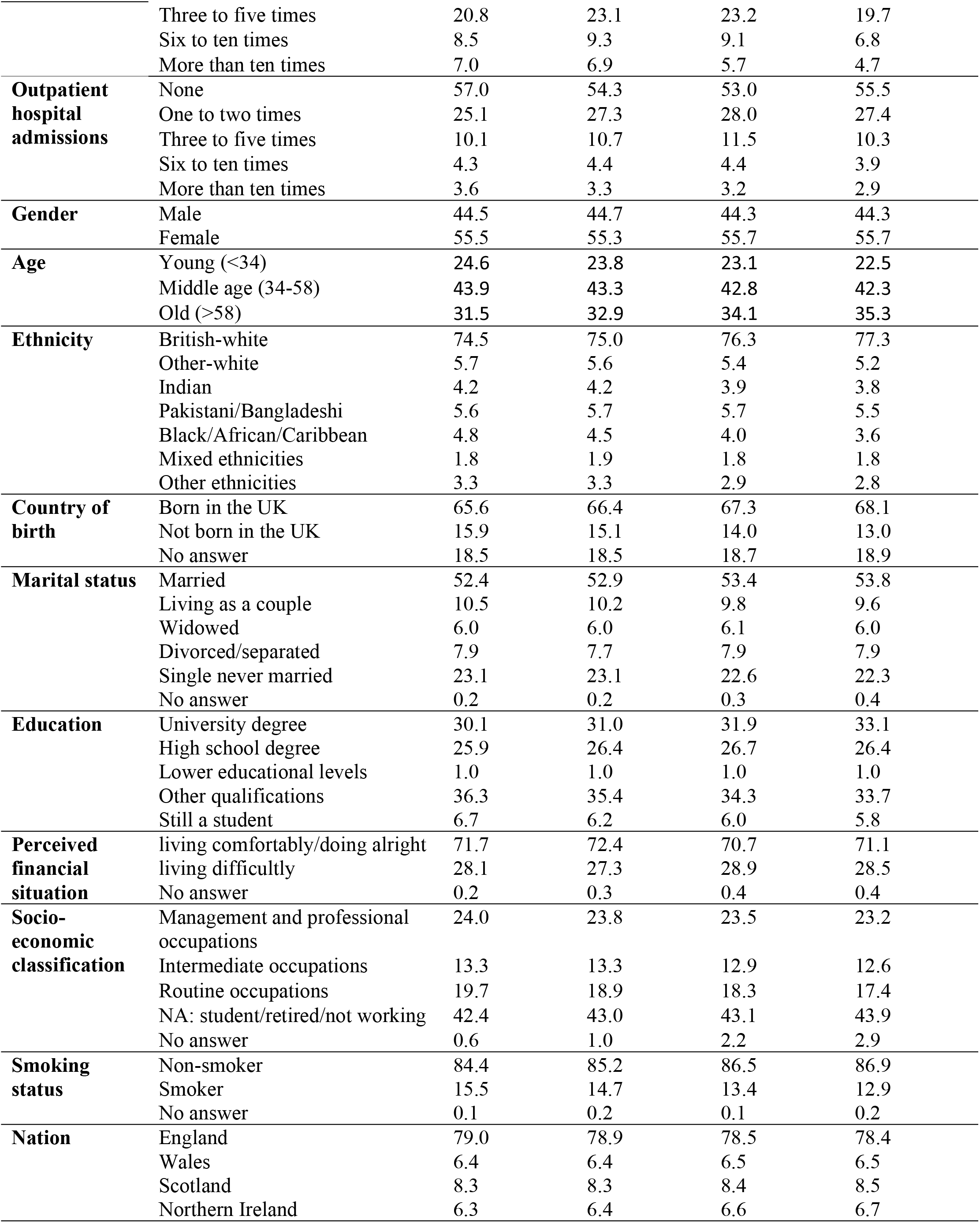
Description of GP visits, outpatient hospital admissions and individual’s socioeconomic and lifestyle covariates for each wave of the UKHLS sample (N=140,466 surveys from 46,442 individuals)

|  |  | **Wave7**<br>**(2015-2017)**<br>**N=38,663** | **Wave8**<br>**(2016-2018)**<br>**N=36,319** | **Wave9**<br>**(2017-2019)**<br>**N=33,727** | **Wave10**<br>**(2018-2019)**<br>**N=31,757** |
| --- | --- | --- | --- | --- | --- |
|  |  | **Percent** | **Percent** | **Percent** | **Percent** |
| **GP visits** | None | 26.7 | 21.1 | 21.2 | 28.3 |
|  | One to two times | 37.0 | 39.5 | 40.8 | 40.6 |
|  | Three to five times | 20.8 | 23.1 | 23.2 | 19.7 |
|  | Six to ten times | 8.5 | 9.3 | 9.1 | 6.8 |
|  | More than ten times | 7.0 | 6.9 | 5.7 | 4.7 |
| **Outpatient hospital admissions** | None | 57.0 | 54.3 | 53.0 | 55.5 |
|  | One to two times | 25.1 | 27.3 | 28.0 | 27.4 |
|  | Three to five times | 10.1 | 10.7 | 11.5 | 10.3 |
|  | Six to ten times | 4.3 | 4.4 | 4.4 | 3.9 |
|  | More than ten times | 3.6 | 3.3 | 3.2 | 2.9 |
| **Gender** | Male | 44.5 | 44.7 | 44.3 | 44.3 |
|  | Female | 55.5 | 55.3 | 55.7 | 55.7 |
| **Age** | Young (<34) | 24.6 | 23.8 | 23.1 | 22.5 |
|  | Middle age (34-58) | 43.9 | 43.3 | 42.8 | 42.3 |
|  | Old (>58) | 31.5 | 32.9 | 34.1 | 35.3 |
| **Ethnicity** | British-white | 74.5 | 75.0 | 76.3 | 77.3 |
|  | Other-white | 5.7 | 5.6 | 5.4 | 5.2 |
|  | Indian | 4.2 | 4.2 | 3.9 | 3.8 |
|  | Pakistani/Bangladeshi | 5.6 | 5.7 | 5.7 | 5.5 |
|  | Black/African/Caribbean | 4.8 | 4.5 | 4.0 | 3.6 |
|  | Mixed ethnicities | 1.8 | 1.9 | 1.8 | 1.8 |
|  | Other ethnicities | 3.3 | 3.3 | 2.9 | 2.8 |
| **Country of birth** | Born in the UK | 65.6 | 66.4 | 67.3 | 68.1 |
|  | Not born in the UK | 15.9 | 15.1 | 14.0 | 13.0 |
|  | No answer | 18.5 | 18.5 | 18.7 | 18.9 |
| **Marital status** | Married | 52.4 | 52.9 | 53.4 | 53.8 |
|  | Living as a couple | 10.5 | 10.2 | 9.8 | 9.6 |
|  | Widowed | 6.0 | 6.0 | 6.1 | 6.0 |
|  | Divorced/separated | 7.9 | 7.7 | 7.9 | 7.9 |
|  | Single never married | 23.1 | 23.1 | 22.6 | 22.3 |
|  | No answer | 0.2 | 0.2 | 0.3 | 0.4 |
| **Education** | University degree | 30.1 | 31.0 | 31.9 | 33.1 |
|  | High school degree | 25.9 | 26.4 | 26.7 | 26.4 |
|  | Lower educational levels | 1.0 | 1.0 | 1.0 | 1.0 |
|  | Other qualifications | 36.3 | 35.4 | 34.3 | 33.7 |
|  | Still a student | 6.7 | 6.2 | 6.0 | 5.8 |
| **Perceived financial situation** | living comfortably/doing alright | 71.7 | 72.4 | 70.7 | 71.1 |
|  | living difficultly | 28.1 | 27.3 | 28.9 | 28.5 |
|  | No answer | 0.2 | 0.3 | 0.4 | 0.4 |
| **Socio-economic classification** | Management and professional occupations | 24.0 | 23.8 | 23.5 | 23.2 |
|  | Intermediate occupations | 13.3 | 13.3 | 12.9 | 12.6 |
|  | Routine occupations | 19.7 | 18.9 | 18.3 | 17.4 |
|  | NA: student/retired/not working | 42.4 | 43.0 | 43.1 | 43.9 |
|  | No answer | 0.6 | 1.0 | 2.2 | 2.9 |
| **Smoking status** | Non-smoker | 84.4 | 85.2 | 86.5 | 86.9 |
|  | Smoker | 15.5 | 14.7 | 13.4 | 12.9 |
|  | No answer | 0.1 | 0.2 | 0.1 | 0.2 |
| **Nation** | England | 79.0 | 78.9 | 78.5 | 78.4 |
|  | Wales | 6.4 | 6.4 | 6.5 | 6.5 |
|  | Scotland | 8.3 | 8.3 | 8.4 | 8.5 |
|  | Northern Ireland | 6.3 | 6.4 | 6.6 | 6.7 |

**Table 2:** Exposure description and correlation matrix of air pollutants (N=42,619 LSOAs; time=2015-2019)

|  | Pearson's correlation coefficient |  |  |  | Mean | SD | Median | Interquartile range |
| --- | --- | --- | --- | --- | --- | --- | --- | --- |
| | NO<sub>2</sub><br>($\mu\text{g}/\text{m}^3$) | SO<sub>2</sub><br>($\mu\text{g}/\text{m}^3$) | PM10<br>($\mu\text{g}/\text{m}^3$) | PM2.5<br>($\mu\text{g}/\text{m}^3$) | | | | |
| NO<sub>2</sub> ($\mu\text{g}/\text{m}^3$) | 1.00 | | | | 14.05 | 6.64 | 13.29 | 8.35 |
| SO<sub>2</sub> ($\mu\text{g}/\text{m}^3$) | 0.55 | 1.00 | | | 1.44 | 0.60 | 1.34 | 0.75 |
| PM10 ($\mu\text{g}/\text{m}^3$) | **0.72** | 0.43 | 1.00 | | 13.37 | 3.04 | 13.56 | 4.58 |
| PM2.5 ($\mu\text{g}/\text{m}^3$) | **0.74** | 0.47 | **0.98** | 1.00 | 8.70 | 2.14 | 8.89 | 3.29 |

Strong correlations with correlation coefficient $\geq 0.70$ are highlighted in bold.

To account for the study design which involves linkages of air pollution to individual-level data at the LSOAs level and to account for the repeated individuals’ responses across time, the association of the GP visits and outpatient hospital admission ordinal outcomes with each of NO_2_, SO_2_, PM10, and PM2.5 pollutants was assessed using three-levels (repeated individual responses across time nested within LSOAs) mixed-effects ordered logistic models. These models were adjusted for the socioeconomic and lifestyle covariates and the year (2015-2019) dummies. In a supplementary analysis, we also demonstrate the association of GP visits and hospital admissions with each of the socioeconomic and lifestyle covariates (Appendix: Tables A1 and A2).

In further analysis, we decomposed the overall effect of air pollution (linked at the LSOAs level) on GP visits and outpatient hospital admissions into *between* (*spatial*) and *within* (*temporal*) effects. *Between* effects were used to determine the *spatial* effect of air pollution by computing the mean of air pollution across the five years of follow-up (2015-2019) for each LSOA. In contrast, *within* effects were used to determine the *temporal* effect of air pollution by calculating the yearly air pollution deviation from the five years mean for each LSOA. Therefore, the multilevel mixed-effects ordered logistic models were used to examine the overall (equation 1) effect of air pollution as well as the *between* and *within* effects (equation 2) of air pollution on GP visits and outpatient hospital admissions.

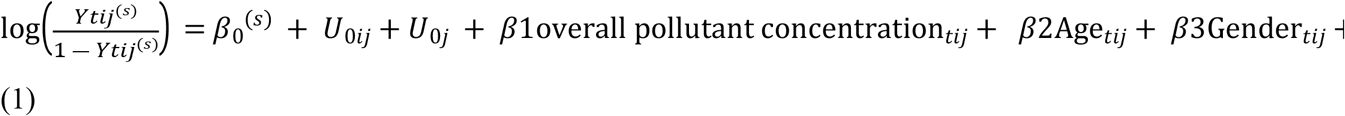

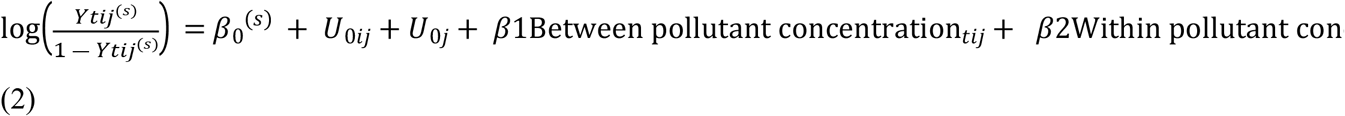

Where *Y*_*tij*_ is the outcome (GP visits or outpatient hospital admissions) for individual *i*, in LSOA *j* at year *t*; s is the level of an ordered category with s levels for GP visits or hospital admission outcomes; *β*_*1*_, *β*_*2*_ …. *β*_*12*_ are the slopes of fixed effects; *β*_*0*_ is the fixed intercept; *U*_*0ij*_ is level 2 random intercept of individuals nested in LSOAs; *U*_*0j*_ is level 3 random intercept of LSOAs;

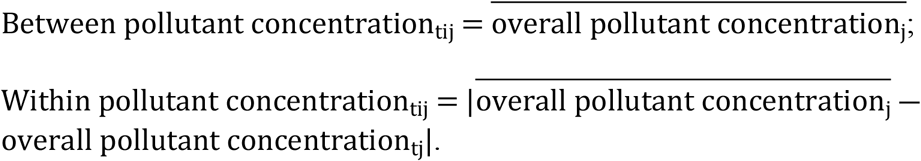

Finally, we incorporated into the multilevel ordered logistic models an interaction term between ethnicity and each of NO_2_, SO_2_, PM10, and PM2.5 pollutants and between country of birth and each of the four pollutants to assess whether the association between air pollution and GP visits and hospital admissions varies between ethnic groups and by country of birth. Interaction terms were incorporated into the overall pollutant models and into the *between-within* models, each at a time. Coefficient plots were used to visualize the interaction analysis odds ratios.

Statistical analysis was conducted using STATA software (StataCorp. 2015. Stata Statistical Software: Release 14. College Station, TX: StataCorp LP) and spatial pre-processing was conducted using ArcGIS Pro software. Regression results were reported in terms of odds ratios (ORs) and 95% confidence intervals (CIs) per 1 μg/m^3^ increase in the air pollutants. Statistical significance was considered at a P-value of less than 0.05.

### 2.4. Ethical considerations

This paper was granted ethical approval on the 14th of May 2020 by the author’s affiliated institution (School of Geography and Sustainable Development Ethics Committee, acting on behalf of the University Teaching and Research Ethics Committee (UTREC) at the University of St Andrews). The paper uses secondary adult (age 16+) fully anonymised data from the “Understanding Society: The UK Household Longitudinal Study (UKHLS)” and authors did not have access to potentially identifying information; thus, obtaining participants’ informed consent is not applicable and was waved by the authors’ institution ethics committee. The University of Essex responsible for the UKHLS data collection and management has already obtained written informed consent from all the study participants (33). Requesting consent for health record linkage was approved at Wave 1 by the National Research Ethics Service (NRES) Oxfordshire REC A (08/H0604/124), and at Wave 4 by NRES Southampton REC A (11/SC/0274). Approval for the collection of biosocial data by trained nurses in Waves 2 and 3 of the main survey was obtained from the National Research Ethics Service (Understanding Society - UK Household Longitudinal Study: A Biosocial Component, Oxfordshire A REC, Reference: 10/H0604/2).

## 3. Results

### 3.1. Description of GP visits, hospital admissions and individuals’ socioeconomic and lifestyle covariates

In all the four waves, most of the individuals visited a GP one to two times, did not go to a hospital as outpatients, were females, aged between 34 and 58 years old, had British-white ethnicity, were born in the UK, were married, had a university degree or other educational qualifications, reported a comfortable/alright financial situation, had a managerial or professional socioeconomic classification (if working), were non-smokers, and lived in England (Table 1).

### 3.2. Description of air pollution

A high correlation (Pearson’s coefficient ≥ 0.7) was observed between NO_2_, PM10, and PM2.5 pollutants, which could be explained by the chemical reactions between particulate matter and NO_2_ pollutants in the atmosphere. Across the five years of follow-up (2015-2019), the mean of NO_2_ was 14.05 μg/m^3^ (SD=6.64), the mean of SO_2_ was 1.44 μg/m^3^ (SD=0.60), the mean of PM10 was 13.37 μg/m^3^ (SD=3.04), and the mean of PM2.5 was 8.70 μg/m^3^ (SD=2.14) (Table2).

### 3.3. The spatial-temporal effect of air pollution on GP visits and hospital admissions

Results showed that higher visits to a GP are associated with higher concentrations of NO_2_ (OR=1.011, 95%CI=1.007-1.015) and SO_2_ (OR=1.123, 95%CI=1.087-1.160) pollutants. Higher odds of outpatient hospital admissions were observed with increased concentrations of NO_2_ (OR=1.009, 95%CI=1.006-1.013), SO_2_ (OR=1.063, 95%CI=1.030-1.097), PM10 (OR=1.013, 95%CI=1.006-1.021) and PM2.5 (OR=1.022, 95%CI=1.012-1.032) pollutants (Table 3).

**Table 3:** The association of air pollution with GP visits and outpatient hospital admissions (N=140,466 surveys from 46,442 individuals)

|  | GP visits | Outpatient hospital admissions |
| --- | --- | --- |
|  | OR [95%CI] | OR [95%CI] |
| **Overall pollution effect** |  |  |
| NO<sub>2</sub> (µg/m<sup>3</sup>) | 1.011 [1.007, 1.015]** | 1.009 [1.006, 1.013]** |
| SO<sub>2</sub> (µg/m<sup>3</sup>) | 1.123 [1.087, 1.160]** | 1.063 [1.030, 1.097]** |
| PM10 (µg/m<sup>3</sup>) | 1.003 [0.995, 1.010] | 1.013 [1.006, 1.021]** |
| PM2.5 (µg/m<sup>3</sup>) | 1.004 [0.994, 1.015] | 1.022 [1.012, 1.032]** |
| **Between pollution effect** |  |  |
| NO<sub>2</sub> (µg/m<sup>3</sup>) | 1.011 [1.007, 1.015]** | 1.010 [1.007, 1.014]** |
| SO<sub>2</sub> (µg/m<sup>3</sup>) | 1.123 [1.077, 1.170]** | 1.053 [1.013, 1.095]* |
| PM10 (µg/m<sup>3</sup>) | 1.005 [0.997, 1.013] | 1.014 [1.006, 1.022]** |
| PM2.5 (µg/m<sup>3</sup>) | 1.009 [0.997, 1.022] | 1.024 [1.012, 1.035]** |
| **Within pollution effect** |  |  |
| NO<sub>2</sub> (µg/m<sup>3</sup>) | 0.999 [0.977, 1.022] | 0.994 [0.970, 1.018] |
| SO<sub>2</sub> (µg/m<sup>3</sup>) | 1.068 [0.972, 1.173] | 1.011 [0.915, 1.118] |
| PM10 (µg/m<sup>3</sup>) | 1.009 [0.982, 1.036] | 1.009 [0.980, 1.038] |
| PM2.5 (µg/m<sup>3</sup>) | 1.000 [0.963, 1.039] | 0.997 [0.958, 1.039] |

\*\*P-value <0.01; \*P-value<0.05
ORs and 95%CI are expressed in terms of 1 µg/m<sup>3</sup> increase in the air pollutants; Models are adjusted for age, gender, ethnicity, country of birth, marital status, education, perceived financial situation, socioeconomic classification, smoking status, and year dummies (2015-2019).

Decomposing the overall effect of air pollution on GP visits and hospital admissions into *between* (spatial: across LSOAs) and *within* (temporal: across years within each LSOA) effects, showed significant positive associations for the *between* effect on GP visits for NO_2_ (OR=1.011, 95%CI=1.007-1.015) and SO_2_ (OR=1.123, 95%CI=1.077, 1.170) pollutants, and a significant *between* effect on outpatient hospital admissions for all the four pollutants. No significant *within* effects were noted for all the four pollutants neither on GP visits nor on hospital admissions (Table 3).

### 3.4. The association of air pollution with GP visits and hospital admissions by ethnicity and country of birth

Examining the association between ethnicity and GP visits revealed higher odds of GP visits among Indian (OR=1.301, 95%CI=1.159-1.462), Pakistani/Bangladeshi (OR=1.525, 95%CI=1.372-1.694), and mixed ethnicities (OR=1.249, 95%CI=1.079-1.447) in comparison to British-white and among non-UK-born individuals (OR=1.239, 95%CI=1.148-1.337) compared to UK-born (Appendix: Table A1). In contrast, higher odds of outpatient hospital admissions were observed only among mixed ethnicities (OR=1.385, 95%CI=1.199-1.600), while Pakistani/Bangladeshi (OR=0.886, 95%CI=0.800-0.981) and non-UK-born individuals (OR=0.893, 95%CI=0.828-0.963) exhibited lower odds of hospital admissions (Appendix: Table A2).

Our analysis showed no differences between ethnic minorities and British-white for the effect of air pollution on GP visits and outpatient hospital admissions (Fig 2 and Fig 3). Thus, ethnic minorities do not seem to experience greater health-related effects from exposure to air pollution compared to the rest of population. The only exception were non-UK-born individuals who were more likely to visit a GP than UK-born individuals with increasing concentrations of all the four pollutants (Fig 2).

**Fig 2:**
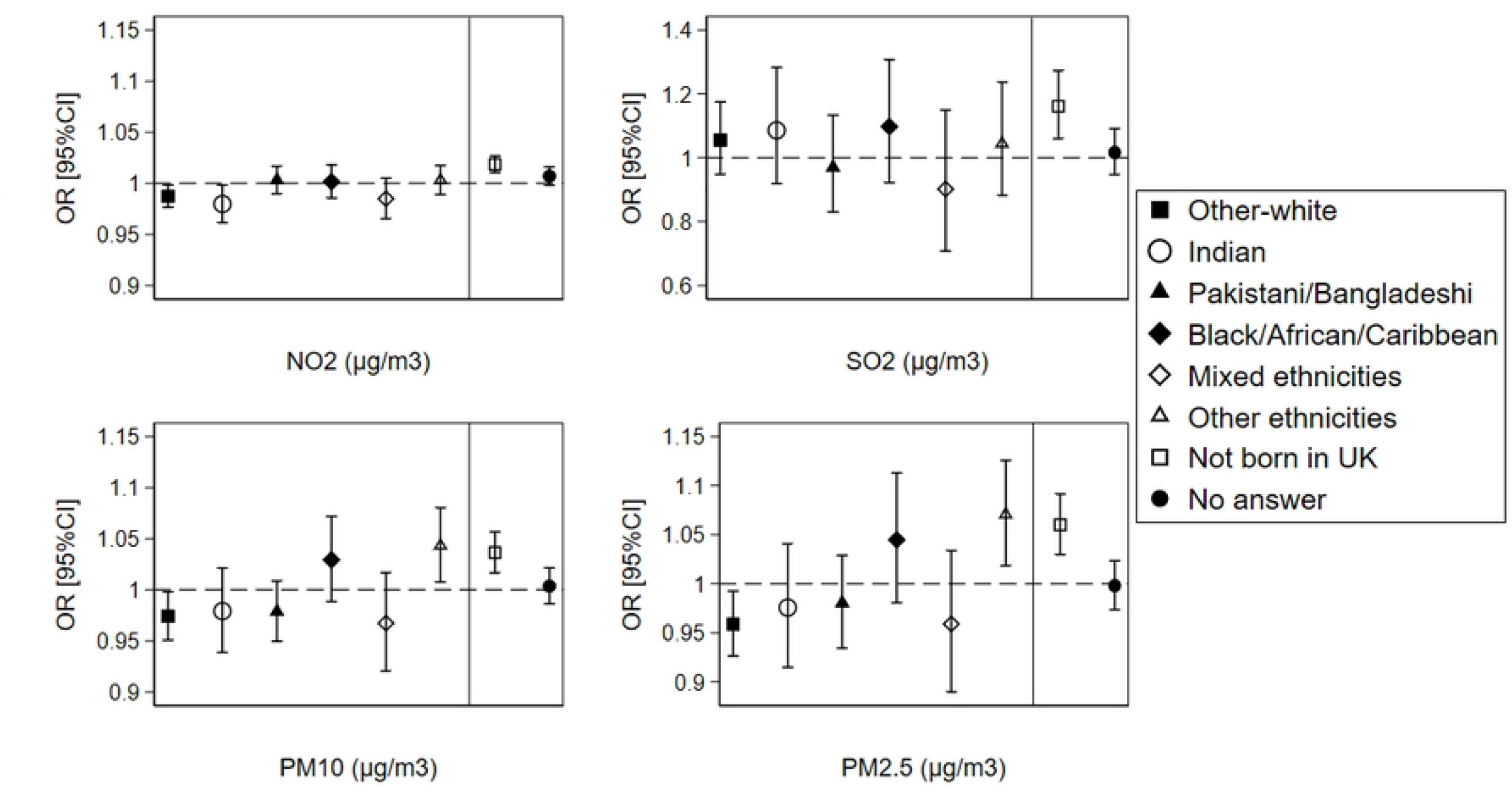
The overall effect of air pollution on GP visits by ethnicity and country of birth (N=140,466 surveys from 46,442 individuals) ORs and 95%CIs are expressed in terms of 1 μg/m^3^ increase in the air pollutants; The dashed line is placed at OR=1 as a cut-off for statistically insignificant results; The solid line separates between the air pollution-ethnicity interaction models and the air pollution-country of birth interaction models; Air pollution-ethnicity interaction models where the reference category is “British-white” are adjusted for country of birth, age, gender, marital status, education, perceived financial situation, socioeconomic classification, smoking status, and year dummies (2015-2019); Air pollution-country of birth interaction models where the reference category is “born in UK” are adjusted for ethnicity, age, gender, marital status, education, perceived financial situation, socioeconomic classification, smoking status, and year dummies (2015-2019).

**Fig 3:**
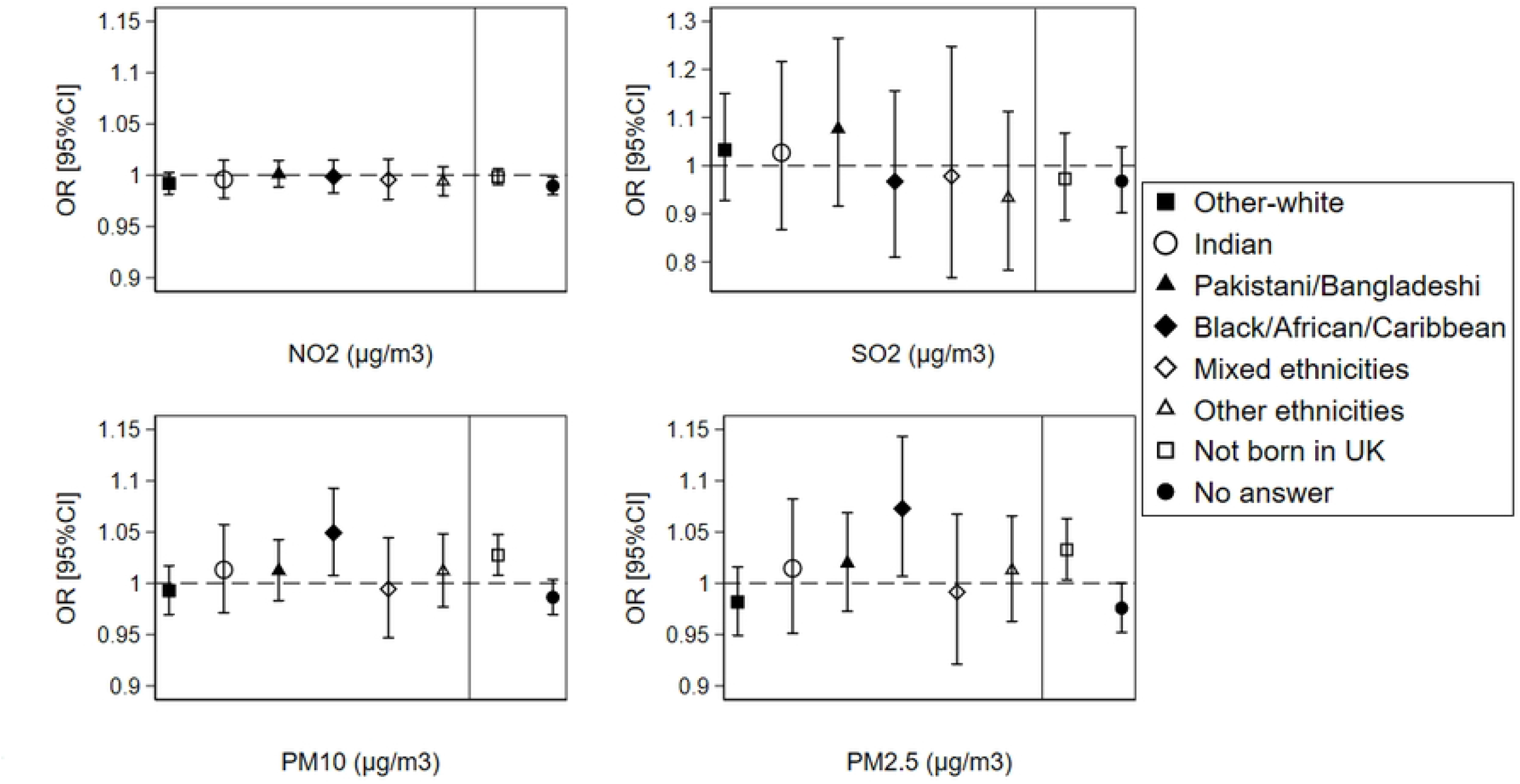
The overall effect of air pollution on outpatient hospital admissions by ethnicity and country of birth (N=140,466 surveys from 46,442 individuals) ORs and 95%CIs are expressed in terms of 1 μg/m^3^ increase in the air pollutants; The dashed line is placed at OR=1 as a cut-off for statistically insignificant results; The solid line separates between the air pollution-ethnicity interaction models and the air pollution-country of birth interaction models; Air pollution-ethnicity interaction models where the reference category is “British-white” are adjusted for country of birth, age, gender, marital status, education, perceived financial situation, socioeconomic classification, smoking status, and year dummies (2015-2019); Air pollution-country of birth interaction models where the reference category is “born in UK” are adjusted for ethnicity, age, gender, marital status, education, perceived financial situation, socioeconomic classification, smoking status, and year dummies (2015-2019).

Analysing the *between-within* (spatial-temporal) effects of air pollution on GP visits and hospital admissions by ethnicity and country of birth did not show significant associations neither for the *between* nor for the *within* effects (Fig 4 and Fig 5); except for non-UK-born individuals who showed higher odds of GP visits with increasing concentrations of all the four pollutants for the *between* (spatial) effects in comparison to UK-born individuals (Fig 4).

**Fig 4:**
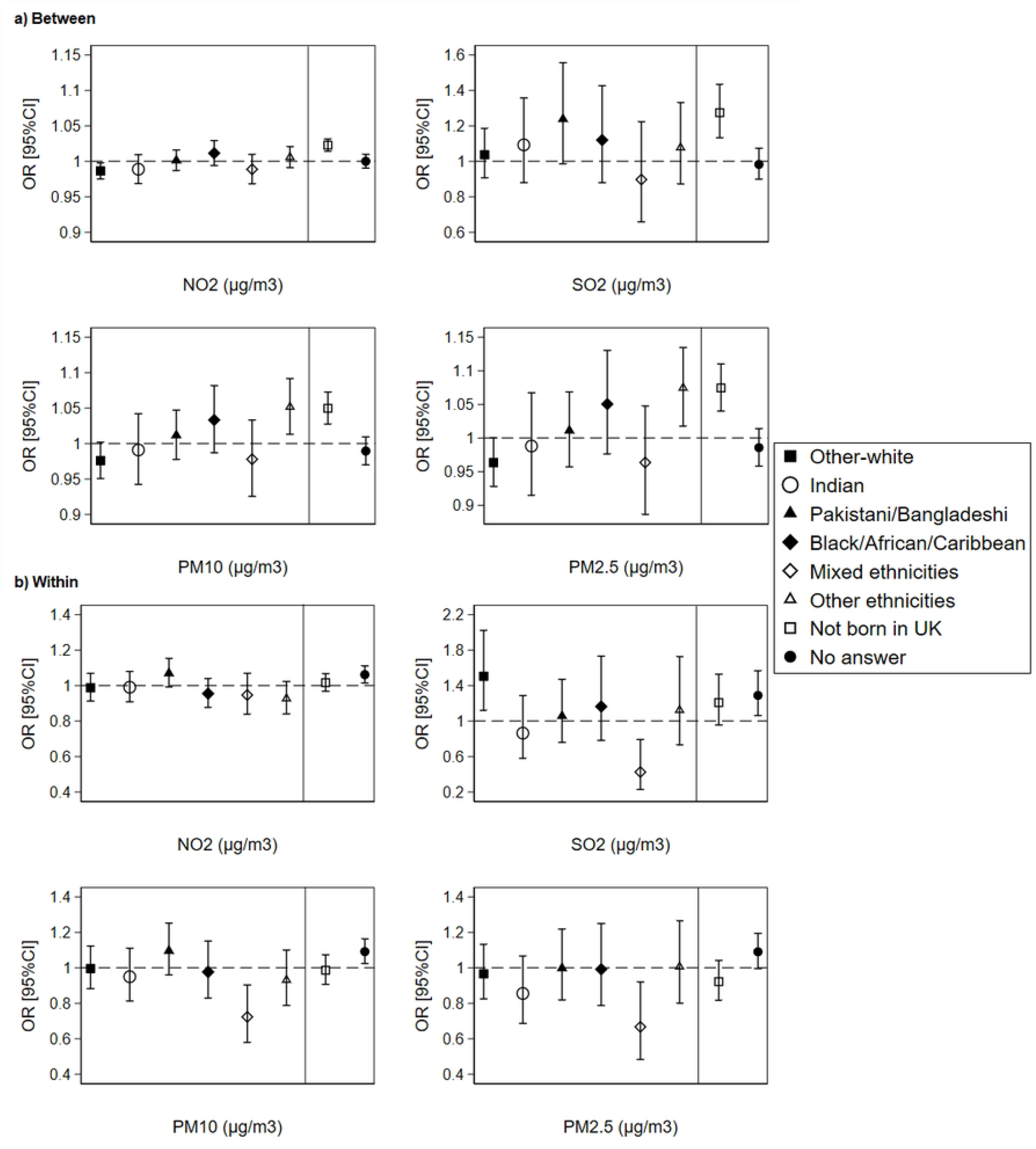
The *between-within* (spatial-temporal) effect of air pollution on GP visits by ethnicity and country of birth (N=140,466 surveys from 46,442 individuals) ORs and 95%CIs are expressed in terms of 1 μg/m^3^ increase in the air pollutants; The dashed line is placed at OR=1 as a cut-off for statistically insignificant results; The solid line separates between the air pollution-ethnicity interaction models and the air pollution-country of birth interaction models; Air pollution-ethnicity interaction models where the reference category is “British-white” are adjusted for country of birth, age, gender, marital status, education, perceived financial situation, socioeconomic classification, smoking status, and year dummies (2015-2019); Air pollution-country of birth interaction models where the reference category is “born in UK” are adjusted for ethnicity, age, gender, marital status, education, perceived financial situation, socioeconomic classification, smoking status, and year dummies (2015-2019).

**Fig 5:**
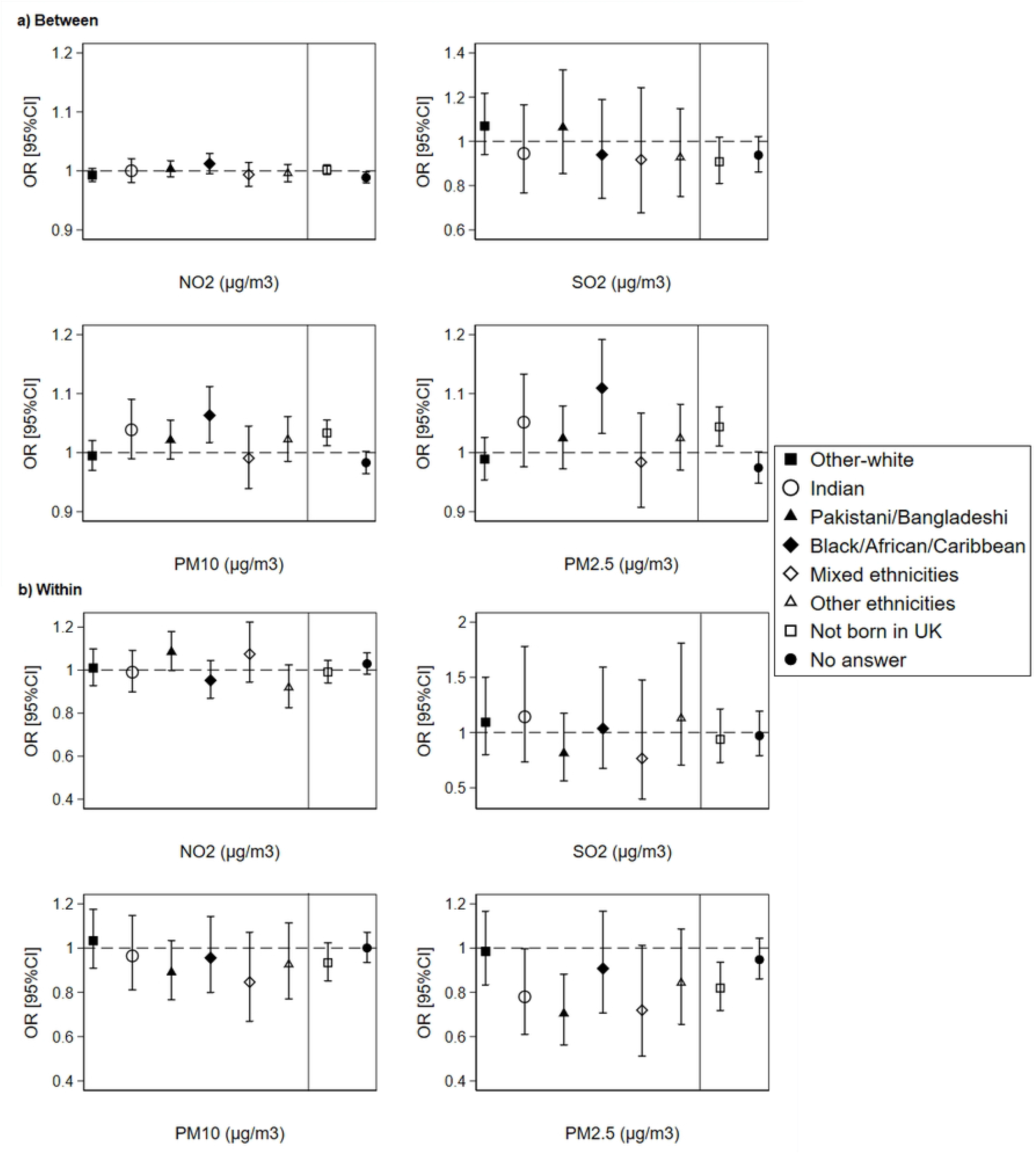
The *between-within* (spatial-temporal) effect of air pollution on outpatient hospital admissions by ethnicity and country of birth (N=140,466 surveys from 46,442 individuals) ORs and 95%CIs are expressed in terms of 1 μg/m^3^ increase in the air pollutants; The dashed line is placed at OR=1 as a cut-off for statistically insignificant results; The solid line separates between the air pollution-ethnicity interaction models and the air pollution-country of birth interaction models; Air pollution-ethnicity interaction models where the reference category is “British-white” are adjusted for country of birth, age, gender, marital status, education, perceived financial situation, socioeconomic classification, smoking status, and year dummies (2015-2019); Air pollution-country of birth interaction models where the reference category is “born in UK” are adjusted for ethnicity, age, gender, marital status, education, perceived financial situation, socioeconomic classification, smoking status, and year dummies (2015-2019).

## 4. Discussion

In this study, we showed that exposure to four air pollutants (NO_2_, SO_2_, PM10, and PM2.5) results in higher frequency of GP visits and outpatient hospital admissions in the UK, which is supported by relevant literature (1, 2, 6, 8, 43, 44). For instance, a 1% increase in the risk of cardiac hospital admissions in Italy was observed per 10 μg/m^3^ increase in PM10 pollutant (45). In Istanbul-Turkey, short-term exposure to NO_2_, PM10, and PM2.5 air pollution was positively associated with increased respiratory hospital admissions between 2013 and 2015 (46). In Australia, a 1% increase in PM10 was associated with 0.6% increase in the number of hospital admissions and 0.5% increase in the number of GP visits (47).

One main novelty of this study is the application of a *between-within* analysis that reveals the *spatial-temporal* effects of air pollution on GP visits and outpatient hospital admissions. Although the *between-within* analysis is widely used in the fields of economics, behavioural finance, and strategic management (32), it is rarely applied in health research (48); and no previous study has assessed the *between-within* effects of air pollution on GP visits and hospital admissions. Applying the *between-within* analysis in this study allowed us to observe significant *between* effects for air pollution on GP visits and hospital admissions; yet, no significant *within* effects were noticed. Therefore, individuals residing in LSOAs with higher exposure to air pollution across the five years of follow-up exhibited higher frequency of GP visits and outpatient hospital admissions than individuals residing in LSOAs with lower pollution exposure. Thus, our study shows strong evidence for the spatial rather than temporal effects of air pollution on GP visits and outpatient hospital admissions in the UK. This could be explained by the emission source of air pollutants and their chemical and physical properties. The major source of NO_2_ emissions is traffic exhaust (49) which varies across both LSOAs (*between: spatial*) and time (*within: temporal*) depending on the number of vehicles and the movement of people. Yet, nitrogen oxides are highly reactive and seasonal pollutants (50), which makes it difficult to capture their temporal variation through yearly measurements. For instance, more NO_2_ will be liberated into the atmosphere during warm seasons due to the chemical reactions between nitrogen oxides and ozone (50). Additionally, NO_2_ is converted into nitric acid by several different reactions in the atmosphere (51). That’s why only spatial (*between*) but not temporal (*within*) effects for NO_2_ pollutant were observed when taking the year as our time measuring unit. On the other hand, industrial processes and power plants are the major sources of SO_2_ pollution (52), which is dominated by spatial (*between*) variation rather than temporal (*within*) variation as building a new factory requires much longer time than purchasing a motor vehicle. Particulate matter results from both traffic exhaust and industrial processes (53), and is considered a more stable pollutant that may stay suspended in the air for long periods of time (51). Given the localized industrial source of particulate matter pollution and its stability in the atmosphere which limits its variation across time, a spatial (*between*) effect on health-related outcomes is expected rather than a temporal (*within*) effect. It should be noted, however, that the insignificant *within* effects for all the pollutants could also be attributed to the relatively short period of follow-up (2015-2019) which does not allow for a high variation in the yearly air pollution concentrations. Thus, for future research, it is recommended to use datasets that allow for a longer follow-up time that might result in significant *within* effects.

Apart from the spatial-temporal (*between-within*) analysis, our study contributed to the topic of ethnic inequalities in health by investigating the differences in the effect of air pollution on GP visits and outpatient hospital admissions by ethnicity and country of birth. Our results showed no differences between ethnic minorities and British-white for the association between air pollution and hospital admissions and GP visits. Nevertheless, non-UK-born individuals were more likely to visit a GP than UK-born individuals with increasing concentrations of all the four pollutants. The more pronounced effect of air pollution on GP visits among non-UK-born individuals compared to UK-born was a spatial one rather than a temporal one as shown in the *between-within* analysis. This shows the importance of contextual location-specific factors in explaining the differences in air pollution exposure and their consequent effect on health-related outcomes for non-UK-born individuals versus UK-born. Research has shown that immigrants (i.e. non-UK-born) often live in large cities and highly populated urbanised regions, near major roads and key transportation networks. This facilitates their movement and increases their chances of personal development, employment, and business start-ups (54).

Furthermore, immigrants often live in low-priced housing, which is often situated in more deprived immigrants’ concentration neighbourhoods or close to major roads (25). These contextual-spatial factors can result in more pronounced mild health consequences and higher frequency of GP visits among immigrants due to the greater exposure to traffic exhaustion and industrial pollution in comparison to UK-born individuals who have additional financial resources and inheritance tenure to move away from metropolitan areas and highly polluted industrial regions. Additionally, immigrants (especially those coming from outside Europe) might have been exposed to higher air pollution concentrations in their countries of origin prior to arriving into the UK, which might result in higher frequency of GP visits due to the cumulative effect of air pollution on health among non-UK-born individuals compared to UK-born.

In addition to highlighting the contributions of the present study, it is equally important to discuss its limitations. First, the study design involved linking individual-level data from the UKHLS dataset to yearly air pollution data at the LSOAs level, under the assumption that individuals residing in the same LSOA are exposed to the same concentration of air pollution. Although LSOAs have high spatial resolution, using another dataset that allows for air pollution linkages at the level of postcodes which possess the highest spatial resolution in the UK is recommended as it would greatly minimize the exposure bias. Another source of exposure bias in this study might come from the assessment of individuals’ exposure to air pollution using the place (LSOA) of residence which does not necessarily equate to the true personal exposure. In reality, an exposure scenario is more complex involving exposure indoors, at the workplace and through commuting patterns. Therefore, future studies are encouraged to integrate air pollution exposure at the residence and workplace and to consider both ambient and indoors air pollution exposure. Finally, our study included individuals followed over five years (2015-2019) and we assessed the association between air pollution and GP visits and hospital admissions at a yearly rather than monthly or daily basis. This could explain the inability of observing significant temporal (*within*) effects of air pollution on GP visits and outpatient hospital admissions due to the low variation in air pollution across the five years of follow-up. Thus, for future research, we recommend using other datasets that allow for longer follow-up time and for monthly or daily air pollution exposure which would increase the temporal and seasonal variation and result in significant temporal effects.

## 5. Conclusion

Using longitudinal individual-level data linked to air pollution data at the LSOAs level, this study supports the presence of a spatial-temporal association between air pollution and higher frequency of GP visits and outpatient hospital admissions in the UK. Results showed stronger *between* (spatial) effects across LSOAs rather than *within* (temporal) effects across time within each LSOA. However, ethnic minorities do not seem to experience greater health-related effects from exposure to air pollution compared to the rest of population. An exception was for non-UK-born individuals who showed higher odds of GP visits with increasing concentrations of air pollution compared to those born in the UK, which is mainly attributed to contextual location-specific differences. Our results are of importance for policymakers toward improving the environmental conditions that would reduce air pollution and eventually lower the frequency of GP visits and outpatient hospital admissions for people residing in the UK.

## Data Availability

We cannot make the data underlying our analysis publicly available due to ethical and legal restrictions. We are using the “Understanding Society: The UK Household Longitudinal Study (UKHLS)” dataset which is an initiative funded by the Economic and Social Research Council and various Government Departments, with scientific leadership by the Institute for Social and Economic Research, University of Essex, and survey delivery by NatCen Social Research and Kantar Public. These data are protected by a copyright license and strictly distributed by the UK Data Service which is the largest digital repository for quantitative and qualitative social science and humanities research data in the UK. Therefore, data underlying our analysis can only be accessed through the UK Data Service for authorized researchers from the following URL: https://beta.ukdataservice.ac.uk/datacatalogue/series/series?id=2000053

## Author contribution statement

Mary Abed Al Ahad: Conceptualization, Investigation, Methodology, Data curation, Formal Analysis, Writing-Original Draft, Writing-Review and Editing, Visualization, Project administration, Funding acquisition.

### Appendix

**Table A1:** The association of GP visits with the socioeconomic and lifestyle covariates (N=140,466 surveys from 46,442 individuals)

|  |  | OR | P-value | lower 95%CI | Upper 95% CI |
| --- | --- | --- | --- | --- | --- |
| Ethnicity | British-white | Ref |  |  |  |
|  | Other-white | 0.840 | 0.001 | 0.762 | 0.927 |
|  | Indian | 1.301 | 0.000 | 1.159 | 1.462 |
|  | Pakistani/Bangladeshi | 1.525 | 0.000 | 1.372 | 1.694 |
|  | Black/African/Caribbean | 1.104 | 0.079 | 0.989 | 1.233 |
|  | Mixed ethnicities | 1.249 | 0.003 | 1.079 | 1.447 |
|  | Other ethnicities | 0.884 | 0.046 | 0.782 | 0.998 |
| **Country of birth** | Born in the UK | Ref |  |  |  |
|  | Not born in the UK | 1.239 | 0.000 | 1.148 | 1.337 |
|  | No answer | 0.775 | 0.000 | 0.725 | 0.829 |
| **Age** | 16-18 | Ref |  |  |  |
|  | 19-23 | 1.112 | 0.027 | 1.012 | 1.222 |
|  | 24-28 | 1.277 | 0.000 | 1.138 | 1.433 |
|  | 29-33 | 1.316 | 0.000 | 1.165 | 1.486 |
|  | 34-38 | 1.202 | 0.003 | 1.064 | 1.358 |
|  | 39-43 | 1.259 | 0.000 | 1.115 | 1.423 |
|  | 44-48 | 1.446 | 0.000 | 1.281 | 1.631 |
|  | 49-53 | 1.615 | 0.000 | 1.431 | 1.824 |
|  | 54-58 | 1.909 | 0.000 | 1.689 | 2.157 |
|  | 59-63 | 1.963 | 0.000 | 1.733 | 2.225 |
|  | 64-68 | 2.084 | 0.000 | 1.834 | 2.367 |
|  | 69-73 | 2.306 | 0.000 | 2.023 | 2.628 |
|  | 74-78 | 2.823 | 0.000 | 2.458 | 3.242 |
|  | >78 | 3.547 | 0.000 | 3.071 | 4.096 |
| **Gender** | Male | Ref |  |  |  |
|  | Female | 2.235 | 0.000 | 2.147 | 2.327 |
| **Education** | University degree | Ref |  |  |  |
|  | High school degree | 1.054 | 0.056 | 0.999 | 1.112 |
|  | Lower educational levels | 1.496 | 0.000 | 1.230 | 1.819 |
|  | Other qualifications | 1.452 | 0.000 | 1.366 | 1.543 |
|  | Still a student | 0.799 | 0.000 | 0.730 | 0.876 |
| **Marital status** | Married | Ref |  |  |  |
|  | Living as a couple | 1.141 | 0.000 | 1.071 | 1.217 |
|  | Widowed | 0.916 | 0.050 | 0.839 | 1.000 |
|  | Divorced/separated | 1.220 | 0.000 | 1.137 | 1.309 |
|  | Single never married | 0.990 | 0.745 | 0.931 | 1.052 |
|  | No answer | 0.913 | 0.461 | 0.716 | 1.164 |
| **Subjective financial situation** | Living comfortably/doing alright | Ref |  |  |  |
|  | Living difficultly | 1.501 | 0.000 | 1.451 | 1.552 |
|  | No answer | 0.819 | 0.093 | 0.648 | 1.034 |
| **Socio-economic classification** | Management and professional occupations | Ref |  |  |  |
|  | Intermediate occupations | 0.992 | 0.791 | 0.935 | 1.053 |
|  | Routine occupations | 1.117 | 0.000 | 1.056 | 1.182 |
|  | NA: student/retired/not working | 1.845 | 0.000 | 1.749 | 1.946 |
|  | No answer | 1.013 | 0.830 | 0.904 | 1.135 |
| **Smoking status** | Non smoker | Ref |  |  |  |
|  | Smoker | 1.128 | 0.000 | 1.072 | 1.187 |
|  | No answer | 0.961 | 0.815 | 0.686 | 1.345 |
| **Time dummies** | 2015 | Ref |  |  |  |
|  | 2016 | 1.151 | 0.000 | 1.109 | 1.194 |
|  | 2017 | 1.199 | 0.000 | 1.155 | 1.244 |
|  | 2018 | 0.890 | 0.000 | 0.857 | 0.925 |
|  | 2019 | 0.707 | 0.000 | 0.673 | 0.743 |

**Table A2:**
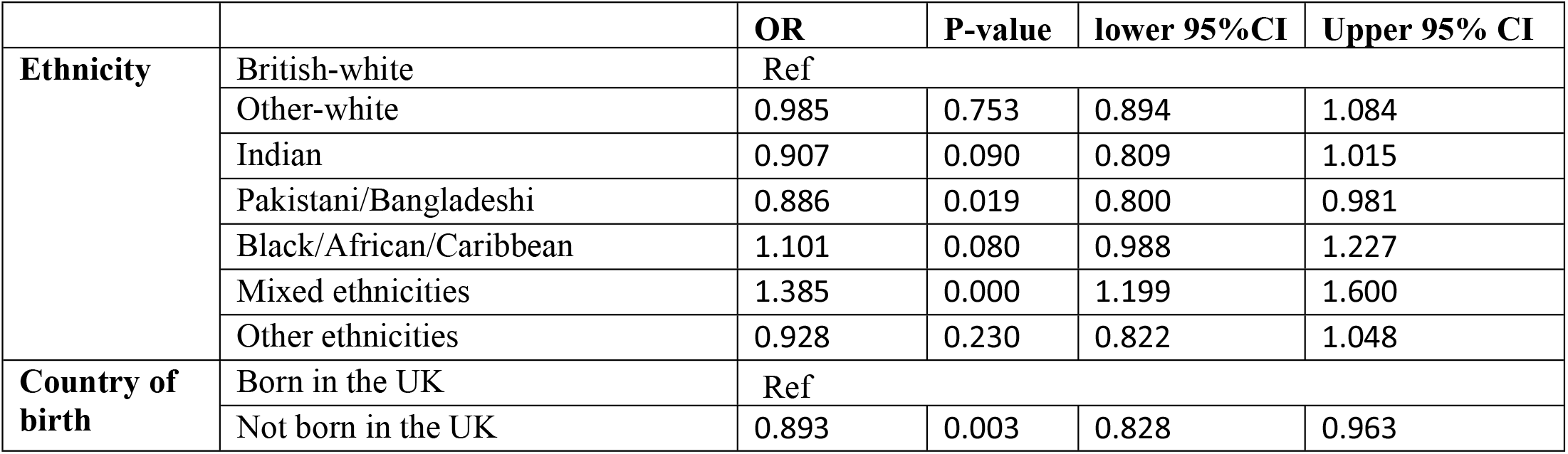
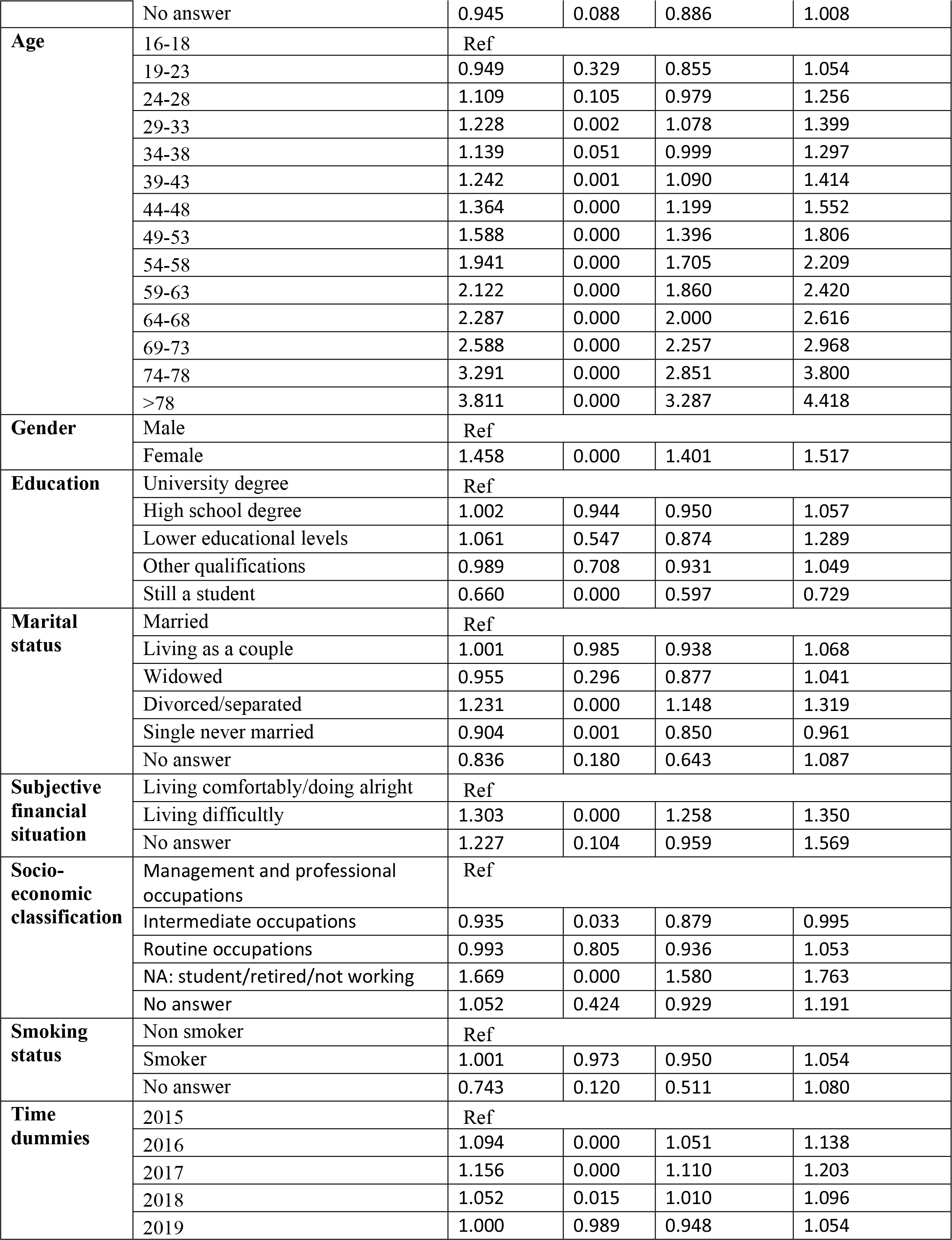
The association of outpatient hospital admissions with the socioeconomic and lifestyle covariates (N=140,466 surveys from 46,442 individuals)

|  |  | OR | P-value | lower 95%CI | Upper 95% CI |
| --- | --- | --- | --- | --- | --- |
| **Ethnicity** | British-white | Ref |  |  |  |
|  | Other-white | 0.985 | 0.753 | 0.894 | 1.084 |
|  | Indian | 0.907 | 0.090 | 0.809 | 1.015 |
|  | Pakistani/Bangladeshi | 0.886 | 0.019 | 0.800 | 0.981 |
|  | Black/African/Caribbean | 1.101 | 0.080 | 0.988 | 1.227 |
|  | Mixed ethnicities | 1.385 | 0.000 | 1.199 | 1.600 |
|  | Other ethnicities | 0.928 | 0.230 | 0.822 | 1.048 |
| **Country of birth** | Born in the UK | Ref |  |  |  |
|  | Not born in the UK | 0.893 | 0.003 | 0.828 | 0.963 |
|  | No answer | 0.945 | 0.088 | 0.886 | 1.008 |
| **Age** | 16-18 | Ref |  |  |  |
|  | 19-23 | 0.949 | 0.329 | 0.855 | 1.054 |
|  | 24-28 | 1.109 | 0.105 | 0.979 | 1.256 |
|  | 29-33 | 1.228 | 0.002 | 1.078 | 1.399 |
|  | 34-38 | 1.139 | 0.051 | 0.999 | 1.297 |
|  | 39-43 | 1.242 | 0.001 | 1.090 | 1.414 |
|  | 44-48 | 1.364 | 0.000 | 1.199 | 1.552 |
|  | 49-53 | 1.588 | 0.000 | 1.396 | 1.806 |
|  | 54-58 | 1.941 | 0.000 | 1.705 | 2.209 |
|  | 59-63 | 2.122 | 0.000 | 1.860 | 2.420 |
|  | 64-68 | 2.287 | 0.000 | 2.000 | 2.616 |
|  | 69-73 | 2.588 | 0.000 | 2.257 | 2.968 |
|  | 74-78 | 3.291 | 0.000 | 2.851 | 3.800 |
|  | >78 | 3.811 | 0.000 | 3.287 | 4.418 |
| **Gender** | Male | Ref |  |  |  |
|  | Female | 1.458 | 0.000 | 1.401 | 1.517 |
| **Education** | University degree | Ref |  |  |  |
|  | High school degree | 1.002 | 0.944 | 0.950 | 1.057 |
|  | Lower educational levels | 1.061 | 0.547 | 0.874 | 1.289 |
|  | Other qualifications | 0.989 | 0.708 | 0.931 | 1.049 |
|  | Still a student | 0.660 | 0.000 | 0.597 | 0.729 |
| **Marital status** | Married | Ref |  |  |  |
|  | Living as a couple | 1.001 | 0.985 | 0.938 | 1.068 |
|  | Widowed | 0.955 | 0.296 | 0.877 | 1.041 |
|  | Divorced/separated | 1.231 | 0.000 | 1.148 | 1.319 |
|  | Single never married | 0.904 | 0.001 | 0.850 | 0.961 |
|  | No answer | 0.836 | 0.180 | 0.643 | 1.087 |
| **Subjective financial situation** | Living comfortably/doing alright | Ref |  |  |  |
|  | Living difficultly | 1.303 | 0.000 | 1.258 | 1.350 |
|  | No answer | 1.227 | 0.104 | 0.959 | 1.569 |
| **Socio-economic classification** | Management and professional occupations | Ref |  |  |  |
|  | Intermediate occupations | 0.935 | 0.033 | 0.879 | 0.995 |
|  | Routine occupations | 0.993 | 0.805 | 0.936 | 1.053 |
|  | NA: student/retired/not working | 1.669 | 0.000 | 1.580 | 1.763 |
|  | No answer | 1.052 | 0.424 | 0.929 | 1.191 |
| **Smoking status** | Non smoker | Ref |  |  |  |
|  | Smoker | 1.001 | 0.973 | 0.950 | 1.054 |
|  | No answer | 0.743 | 0.120 | 0.511 | 1.080 |
| **Time dummies** | 2015 | Ref |  |  |  |
|  | 2016 | 1.094 | 0.000 | 1.051 | 1.138 |
|  | 2017 | 1.156 | 0.000 | 1.110 | 1.203 |
|  | 2018 | 1.052 | 0.015 | 1.010 | 1.096 |
|  | 2019 | 1.000 | 0.989 | 0.948 | 1.054 |

